# Health worker compliance with severe malaria treatment guidelines in the context of implementing pre-referral rectal artesunate: an operational study in three high burden countries

**DOI:** 10.1101/2021.11.26.21266917

**Authors:** Aita Signorell, Phyllis Awor, Jean Okitawutshu, Antoinette Tshefu, Elizabeth Omoluabi, Manuel W. Hetzel, Prosciova Athieno, Joseph Kimera, Gloria Tumukunde, Irene Angiro, Jean-Claude Kalenga, Babatunde Akano, Kazeem Ayodeji, Charles Okon, Ocheche Yusuf, Giulia Delvento, Tristan T. Lee, Nina C. Brunner, Mark Lambiris, James Okuma, Nadja Cereghetti, Valentina Buj, Theodoor Visser, Harriet G Napier, Christian Lengeler, Christian Burri

**Author notes:** Equal contribution (Christian Lengeler, Christian Burri).

## Abstract

**Background:** Appropriate clinical management of severe malaria is critical to avert morbidity and death. Recommended treatment consists of an injectable antimalarial followed by a full course of oral artemisinin-based combination therapy (ACT). Children who cannot access prompt parenteral treatment should be administered a single dose of rectal artesunate (RAS) and promptly referred to an appropriate facility for further care. This study aimed to assess compliance with the treatment recommendation in children under 5 years diagnosed with severe malaria and admitted to referral facilities in 3 high-burden sub-Saharan African countries.

**Methods and Findings:** This study accompanied the implementation of RAS as a pre-referral treatment in the Democratic Republic of the Congo (DRC), Nigeria and Uganda. Children under 5 who were admitted at a referral health facility (RHF) with a diagnosis of severe malaria were included. Type and dosage of antimalarial treatment at RHFs was assessed for children referred from a community-based provider and those directly attending the RHF. We used multivariable regression models to assess factors associated with administration of compliant treatment.

RHF data of 7,983 children was analysed for compliance with regards to antimalarials, a subsample of 3,449 children was assessed in more detail for schedule and dosage compliance and method of ACT provision. Overall, 42.0% (3,356/7,983) of admitted children were administered full treatment consisting of a parenteral antimalarial and an ACT, with large variation among study countries (2.7% in Nigeria, 44.5% in Uganda and 50.3% in DRC). Children receiving RAS from a community-based provider were more likely to be administered compliant post-referral medication at RHFs in DRC (adjusted odds ratio (aOR)=2.19, 95% CI 1.60-2.99), but less likely in Uganda (aOR = 0.43, 95% CI 0.19-0.96). Use of injectable antimalarials was very high in all three countries (99.2% (1,344/1,355) in Uganda, 98.1% (413/421) in Nigeria and 94.4% (1,580/1,673) in DRC), with most children receiving the recommended minimum of three doses (99.0% (1,331/1,344) in Uganda, 95.5% (1,509/1,580) in DRC and 92.0% (380/413) in Nigeria). Rather than being administered in the RHF, ACTs were often prescribed at discharge in Nigeria (54.4%, 229/421) and Uganda (53.0%, 715/1,349), while this was rarely done in DRC (0.8%, 14/1,669) where inpatient administration was more common.

**Conclusions:** Directly observed treatment with both a parenteral antimalarial and an ACT was rare and variable between countries, bearing a high risk for incomplete parasite clearance and disease recrudescence. Parenteral artesunate not followed up with a full course of oral ACT constitutes an artemisinin monotherapy and may favour the selection or development of resistant parasites. Stricter health worker compliance with the WHO severe malaria treatment guidelines is therefore needed to effectively manage this disease and further reduce child mortality.

## Introduction

Malaria deaths result from progression of uncomplicated malaria to severe disease (1). The risk of dying from malaria is highest within the first 24 hours after onset of severe symptoms (2). Prompt initiation of treatment is therefore vital to avert severe morbidity and death. The World Health organization (WHO) recommends treatment for severe malaria consists of an injectable antimalarial (artesunate, artemether or quinine) followed by a full course of oral artemisinin-based combination therapy (ACT; (3)).

Despite these very effective and safe treatment options, many children still die from severe malaria. Two main reasons may be responsible for a fatal outcome: firstly, in several endemic countries, a large proportion of children suffering from severe disease lack prompt access to quality health services and never or only belatedly reach the formal health system (4-6). Secondly, the quality of care that a severely ill child receives is often poor (7-10).

To increase access to essential treatments for children, the WHO 2015 malaria treatment guidelines (3) advise that in situations where parenteral antimalarial treatment cannot be administered, a single pre-referral dose of rectal artesunate (RAS) should be given and the patient should be referred to a health facility where injectable treatment is available (based on evidence from a multi-center clinical trial conducted in Bangladesh, Ghana and Tanzania in 2009 (11)). After the WHO prequalification of two RAS products in 2018 (12), endemic countries started to scale up RAS distribution through existing integrated community case management (iCCM) and integrated management of childhood illnesses (IMCI) programs (13). However, little evidence is available regarding the operational feasibility of incorporating RAS into the continuum of care for severe malaria, and the unanticipated consequences this could have on the overall management of this disease. In addition, it is unclear how much impact the introduction of RAS would have under real-world circumstances (14).

The Community Access to Rectal Artesunate for Malaria (CARAMAL) project was designed as a large-scale operational study to explore the feasibility and effectiveness of implementing RAS as a pre-referral treatment (Lengeler, Burri et al., manuscript submitted). The study aimed to assess healthcare seeking patterns (15), RAS use and acceptance (Kimera et al., manuscript in preparation), anti-malarial treatment received at the various points of contact with the health system, health outcome at day 28 (16), as well as health system costs associated with the roll-out of pre-referral RAS (Lambiris et al., manuscript submitted).

RAS on its own is insufficient to cure a severe malaria episode. RAS can hence only be integrated as an effective component of the continuum of care if the management of severely ill children is adequate. It is therefore important to understand the care and treatment that patients receive in health facilities, and whether these are in line with current recommendations. Prompt initiation of parenteral treatment is critical as hospital deaths occur most often within 24 hours of admission and before the antimalarial treatment takes effect (2, 17). Compliance with the WHO guidelines for severe malaria treatment is also important to avoid artemisinin-based monotherapy and the risk of artemisinin-resistance (18).

This paper describes severe malaria treatment provided to children below five years of age in referral health facilities in the context of RAS roll-out and provides evidence for necessary improvements in severe malaria case management in addition to providing this gap-filling commodity.

## Methods

### Study design and study setting

This study was part of the CARAMAL project, a multi-country observational study on the implementation of quality assured pre-referral RAS via community health workers implementing iCCM algorithms, and primary health centres implementing IMCI guidelines. The details of the design and methods of the CARAMAL project have been published elsewhere (Lengeler, Burri et al., manuscript submitted). In short, CARAMAL was designed as a pre-post intervention study which started in April 2018, before the roll-out of RAS by the United Nations Children’s Fund (Unicef) and local health authorities. The post-RAS introduction period went from April/May 2019 until August 2020. The study areas included three health zones in the Democratic Republic of the Congo (DRC; Ipamu, Kenge and Kingandu), three Local Government Areas (LGA) of Adamawa State in Nigeria (Fufore, Mayo-Belwa and Song), and three districts in Uganda (Kole, Kwania and Oyam). These areas were selected based on their malaria incidence rates, the presence of community health workers implementing iCCM, geographic accessibility and safety considerations (Lengeler, Burri et al., manuscript in preparation).

### Participants and procedures

#### Study inclusion

The main data collection component of the CARAMAL study consisted of a patient surveillance system (PSS) in which children under five years with severe febrile illness / suspected severe malaria were provisionally enrolled upon their first contact with community health workers (CHW) implementing iCCM, or primary health centres (PHC) implementing IMCI. Inclusion criteria were aligned with the criteria for administering RAS according to the country guidelines and included age under five years, history of fever plus at least one danger sign defining a severe febrile illness episode according to the national iCCM guidelines (not able to drink or feed anything, unusually sleepy or unconscious, convulsions, vomits everything). Following provisional enrolment of an eligible patient, basic information on inclusion criteria, RAS administration and referral was transmitted to the study team by the healthcare worker according to country specific notification routes, and captured in electronic study forms and registers.

For the present study, patients recruited at community level were excluded from the analysis, if the diagnosis of severe malaria was not confirmed at the referral health facility (RHF). For a comprehensive assessment of severe malaria treatment at referral facilities, we also included children below the age of five directly attending one of the designated RHFs and diagnosed with severe malaria.

To document the case management of admitted children, trained study staff was based at each of the 25 RHFs within the study areas. All monitored RHFs were public or private but not-for-profit institutions, including health center level IV (HC IV) and hospitals in Uganda, cottage hospitals in Nigeria and referral health centers and general reference hospitals in DRC.

All patients were followed up at their homes 28 days after (provisional) enrolment by a community health worker, PHC or RHF. In a structured interview with the child’s caregiver, the child’s current health status, care seeking patterns during the past illness, and antimalarial treatment, including RAS, was recorded. If the patient was found to be deceased on Day 28, the interview was conducted at least 4 weeks after the child’s passing away, to respect the mourning period.

#### Data collection

Case management information was extracted from patients’ hospital records by the trained study staff, and complemented by direct observation and information obtained from resident hospital staff. The main variables extracted included age, sex, weight, dates and times of admission and discharge, clinical assessments and laboratory tests with their results, provisional and final diagnoses, treatment provided throughout the patient’s admission, and antimalarial prescription / dispensing at discharge. At discharge, the health status, as well as any other diagnoses made and the payment scheme for hospitalisation and any drugs were recorded (artesunate and ACT are supposed to be free of charge). Inpatient treatment data was transcribed in real-time on paper forms and then copied onto tablets using structured electronic forms in ODK Collect (https://opendatakit.org/). An updated data collection form implemented two (Nigeria) to four (DRC, Uganda) months after the roll-out of RAS also included dates and times of antimalarial therapy administration, drug type, route, frequency and duration of administration and details of antimalarial prescription / dispensing at discharge. Information on the use of pre-referral RAS was obtained and consolidated from different data sources through the different points of contact with the healthcare system (CHW, PHC, RHF) and from the caregiver’s interview on day 28.

### Definitions

Table S1 shows the recommended first-line and alternative treatments and the recommended dosing as per WHO treatment guidelines (3) as well as the respective country guidelines. In all three countries, severe malaria is supposed to be managed at RHFs with inpatient facilities.

Compliance was assessed along two dimensions: i) compliance with WHO treatment recommendations, i.e. administration of the recommended antimalarial drugs and ii) compliance with the WHO dose recommendations, i.e. prescription of the recommended dose and frequency of antimalarial medication. Treatment compliance was defined as health worker administration of at least one dose of an injectable antimalarial (artesunate, artemether or quinine) and at least one dose of an ACT (artemether-lumefantrine (ALU) or artesunate amodiaquine (ASAQ)) while the patient was still hospitalised. This indicator was computed for the entire study population. The updated data collection form implemented in the post-RAS phase allowed us to refine this assessment and include the number of doses of injectable treatment and the prescription of ACTs at discharge. This allowed us to distinguish between compliant treatment administration (defined as at least three doses of an injectable antimalarial followed by at least one dose of an ACT administered by health workers during hospitalisation) and, more generally, compliant treatment prescription (defined as at least three doses of an injectable antimalarial followed by at least one dose of an ACT administered by a health worker or prescribed or dispensed at RHF discharge).

The second compliance dimension, dosing compliance of parenteral antimalarial treatment and ACTs, was also estimated for the sub-population and was based on the total amount of drug that patients were prescribed. Details on the evaluation of correct injectable and ACT dosing can be found in the Supplementary materials S5. Since ALU and ASAQ were much more commonly used at the study sites than dihydroartemisinin-piperaquine (DHA-PQ), the latter was not included in the analysis.

### Data management and statistical analysis

The fraction of children with a severe malaria diagnosis who received parenteral artesunate, artemether or quinine, and oral ACT was computed. Antimalarial treatment compliance was calculated as proportion of the total sample of included children (N = 7,983), compliant treatment administration and prescription as well as dosing compliance were estimated on a post-RAS subsample only (N = 3,449, updated data collection form).

Age, sex, weight, pre-referral RAS administration, treatment seeking and malaria test result were considered as potential patient-level predictors of compliant treatment. The provider-level predictor analysed was whether hospitalisation or drugs were payable by caregivers; contextual predictors comprised study country, study area (health zone, district or LGA) and seasonality. Potential predictors for treatment compliance were determined by a logistic regression model with a binary outcome variable equal to 1 if treatment was in accordance to WHO recommendations (i.e. injectable antimalarial followed by an ACT). We report a multivariable model adjusting for all other predictors. For weight, which had 12.4% missing values in the total sample, we used a category of missing indicators in order to include all participants in the models.

Results were stratified by country and enrolment location: patients enrolled by a peripheral healthcare provider and successfully completing referral to a RHF, versus patients directly attending a RHF.

For all statistical tests, a *p*-value < 0.05 was considered significant. All analyses were performed using Stata/MP 16.1 (StataCorp, College Station, Texas, USA).

### Ethics statement

The CARAMAL study protocol was approved by the Research Ethics Review Committee of the World Health Organization (WHO ERC, No. ERC.0003008), the Ethics Committee of the University of Kinshasa School of Public Health (No. 012/2018), the Health Research Ethics Committee of the Adamawa State Ministry of Health (S/MoH/1131/I), the National Health Research Ethics Committee of Nigeria (NHREC/01/01/2007-05/05/2018), the Higher Degrees, Research and Ethics Committee of the Makerere University School of Public Health (No. 548), the Uganda National Council for Science and Technology (UNCST, No. SS 4534), and the Scientific and Ethical Review Committee of CHAI (No. 112, 21 Nov 2017). The study is registered on ClinicalTrials.gov (NCT03568344). Only patients whose caregivers provided written informed consent were enrolled in the study.

## Results

### Characteristics of patients

Between April 2018 and July 2020, 14,911 children were provisionally enrolled into the PSS. Caregivers did not provide consent for 1,153 of them. Of the enrolled 13,758 children, 5,656 were not admitted to one of the RHFs monitored by the study team, and 119 did not have a diagnosis of severe malaria at admission. Hence, 7,983 children were included in this analysis (Figure 1). For 3,449 (43.2%) of them, more detailed case management data was recorded with an updated data collection form.

**Figure 1:**
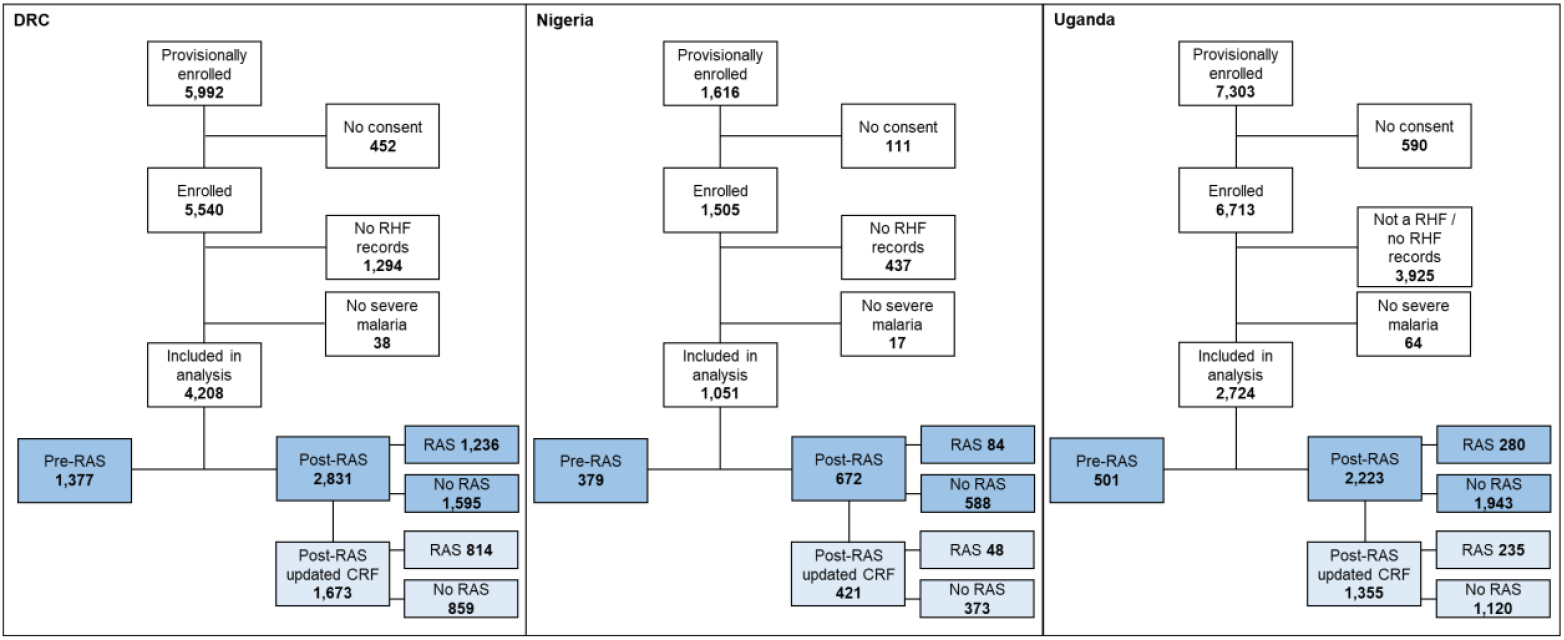
Details of analysis dataset and definitions of compliance dimensions. CRF: data collection form; DRC: Democratic Republic of the Congo; RHF: Referral Health Facility; RAS: Rectal Artesunate

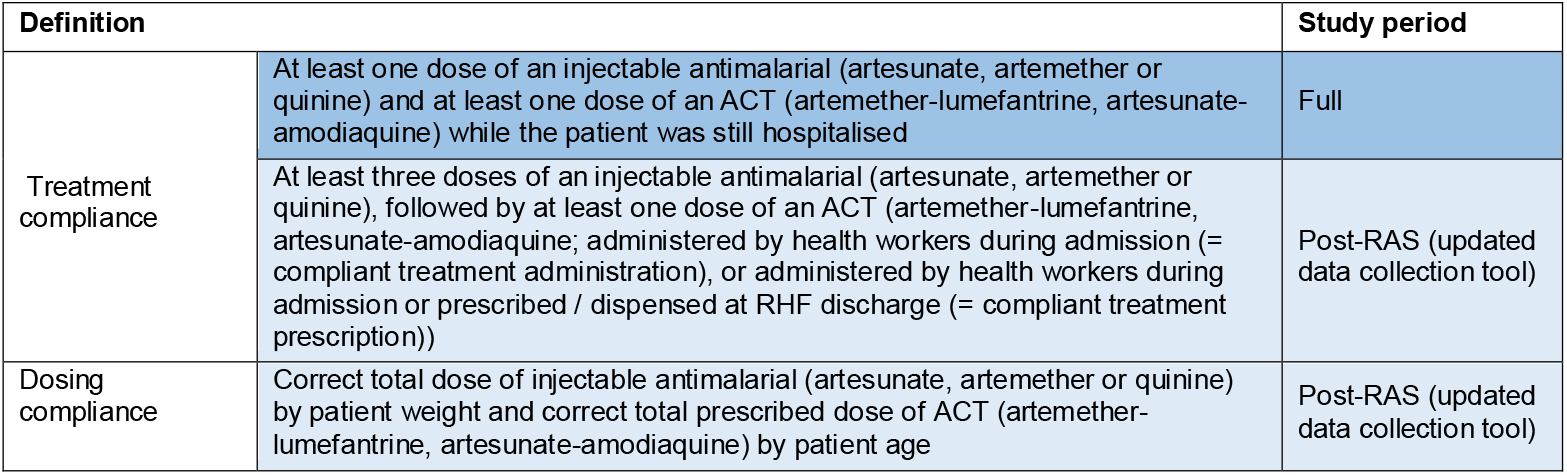

Table 1 (full sample) and Table S2 (sub-sample) show baseline characteristics of the children included in this analysis. 2,381 (29.8%) children undergoing treatment at a RHF were enrolled by a community-based healthcare provider (CHW, PHC) and 5,602 (70.2%) were enrolled directly at a RHF. The median age was 2 years (IQR = 2) and the median weight was 10 kg (IQR = 4). 45.8% were female (42.2% in Nigeria, 47.2% in DRC and 45.1% in Uganda). During the post-RAS implementation period, among the children enrolled by a community-based provider, the proportion who received a dose of pre-referral RAS was higher in DRC (overall 82.6%) and Uganda (73.2%) than in Nigeria (44.2%). Overall, 88.8% of the patients enrolled at a community-based provider and 93.3% of the patients enrolled directly at the RHF had a positive mRDT or microscopy result at some point during their hospital stay.

**Table 1:** Summary characteristics of surveyed patients and exposure variables. RHF: Referral Health Facility; RAS: Rectal Artesunate.

|  | Pooled (3 countries) |  | DRC |  | Nigeria |  | Uganda |  |
| --- | --- | --- | --- | --- | --- | --- | --- | --- |
|  | Community enrolments<br>N = 2,381<br>n (%) | RHF enrolments<br>N = 5,602<br>n (%) | Community enrolments<br>N = 1,939<br>n (%) | RHF enrolments<br>N = 2,269<br>n (%) | Community enrolments<br>N = 217<br>n (%) | RHF enrolments<br>N = 834<br>n (%) | Community enrolments<br>N = 225<br>n (%) | RHF enrolments<br>N = 2,449<br>n (%) |
| **Age (years)** |  |  |  |  |  |  |  |  |
| < 1 | 493 (20.7) | 1061 (18.9) | 428 (22.1) | 475 (20.9) | 19 (8.8) | 91 (10.9) | 46 (20.4) | 495 (19.8) |
| 1 - 2 | 1266 (53.2) | 2966 (53.0) | 996 (51.4) | 1115 (49.1) | 134 (61.8) | 474 (56.8) | 136 (60.4) | 1377 (55.1) |
| 3 - < 5 | 622 (26.1) | 1575 (28.1) | 515 (26.6) | 679 (29.9) | 64 (29.5) | 269 (32.3) | 43 (19.1) | 627 (25.1) |
| **Sex** |  |  |  |  |  |  |  |  |
| Female | 1113 (46.8) | 2542 (45.4) | 931 (48.0) | 1053 (46.4) | 76 (35.0) | 367 (44.0) | 106 (47.1) | 1122 (44.9) |
| **Weight (kg)** |  |  |  |  |  |  |  |  |
| < 8 | 385 (17.4) | 771 (16.2) | 331 (17.2) | 306 (13.7) | 30 (20.0) | 189 (28.0) | 24 (16.6) | 276 (14.8) |
| 8 – 10 | 1068 (48.2) | 2046 (42.9) | 941 (49.0) | 1063 (47.5) | 63 (42.0) | 283 (42.0) | 64 (44.1) | 700 (37.6) |
| > 10 | 764 (34.5) | 1958 (41.0) | 650 (33.8) | 869 (38.8) | 57 (38.0) | 202 (30.0) | 57 (39.3) | 887 (47.6) |
| missing | 164 (6.9) | 827 (14.8) | 17 (0.9) | 31 (1.4) | 67 (30.9) | 160 (19.2) | 80 (35.6) | 636 (25.5) |
| **RAS implementation period & pre-referral RAS use** |  |  |  |  |  |  |  |  |
| Pre-RAS | 594 (25.0) | 1663 (29.7) | 501 (25.8) | 876 (38.6) | 36 (16.6) | 343 (41.1) | 57 (25.3) | 444 (17.8) |
| RAS use: yes | 5 (0.8) | 8 (0.5) | 3 (0.6) | 2 (0.2) | 0 (0.0) | 0 (0.0) | 2 (3.5) | 6 (1.4) |
| RAS use: no | 589 (99.2) | 1655 (99.5) | 498 (99.4) | 874 (99.8) | 36 (100.0) | 343 (100.0) | 55 (96.5) | 438 (98.7) |
| Post-RAS | 1787 (75.1) | 3939 (70.3) | 1438 (74.2) | 1393 (61.4) | 181 (83.4) | 491 (58.9) | 168 (74.7) | 2055 (82.2) |
| RAS use: yes | 1391 (77.8) | 209 (5.3) | 1188 (82.6) | 48 (3.5) | 80 (44.2) | 4 (0.8) | 123 (73.2) | 157 (7.6) |
| RAS use: no | 396 (22.2) | 3730 (94.7) | 250 (17.4) | 1345 (96.6) | 101 (55.8) | 487 (99.2) | 45 (26.8) | 1898 (92.4) |
| **Malaria test\*\*** |  |  |  |  |  |  |  |  |
| positive (mRDT or blood slide) | 2114 (88.8) | 5224 (93.3) | 1695 (87.4) | 2007 (88.5) | 197 (90.8) | 740 (88.7) | 222 (98.7) | 2477 (99.1) |
| negative / not done | 267 (11.2) | 378 (6.8) | 244 (12.6) | 262 (11.6) | 20 (9.2) | 94 (11.3) | 3 (1.3) | 22 (0.9) |
| **Rainy season<sup>oo</sup>** | 1411 (59.3) | 3383 (60.4) | 1096 (56.5) | 1113 (49.1) | 159 (73.3) | 547 (65.6) | 156 (69.3) | 1723 (69.0) |
| **Drugs payable** | 1042 (45.7) | 1967 (39.0) | 863 (46.5) | 1005 (54.3) | 144 (72.0) | 543 (78.1) | 35 (15.6) | 419 (16.8) |
| missing | 101 (4.2) | 557 (9.9) | 84 (4.3) | 418 (18.4) | 17 (7.8) | 139 (16.7) | 0 (0.0) | 0 (0.0) |
| **Hospitalisation payable** | 917 (40.2) | 1329 (26.3) | 874 (47.1) | 859 (46.4) | 33 (16.5) | 76 (10.9) | 10 (4.4) | 394 (15.8) |
| missing | 101 (4.2) | 557 (9.9) | 84 (4.3) | 418 (18.4) | 17 (7.8) | 139 (16.7) | 0 (0.0) | 0 (0.0) |
| **Health Zone / District / LGA** |  |  |  |  |  |  |  |  |
| Kenge / Fufore / Kole | 617 (25.9) | 883 (15.8) | 617 (31.8) | 883 (38.9) | 71 (32.7) | 309 (37.1) | 74 (32.9) | 618 (24.7) |
| Kingandu / Mayo Belwa / Oyam | 503 (21.1) | 278 (5.0) | 503 (25.9) | 278 (12.3) | 123 (56.7) | 227 (27.2) | 119 (52.9) | 1399 (56.0) |
| Ipamu / Song / Apac | 819 (34.4) | 1108 (19.8) | 819 (42.2) | 1108 (48.8) | 23 (10.6) | 298 (35.7) | 32 (14.2) | 482 (19.3) |

Number and column % of those with non-missing data, pooled, by country and by enrolment location; missing data rows are number and column %
\*\* Severe malaria diagnosis was based on clinical assessment, diagnostic test result may or may not have been considered for the diagnosis
<sup>o</sup> WHO general danger signs
<sup>oo</sup> At time of admission; DRC: October – April; Nigeria: May – October; Uganda: April – October

While payment for hospitalisation and drugs was rather frequent in DRC (46.8% paid for hospitalisation, 50.4% for drugs), few caregivers in Uganda did have to pay for anything (14.8% and 16.7% were charged for admission and drugs, respectively). In Nigeria, only 12.2% incurred costs for hospitalisation, but 76.8% were charged for drugs.

### Antimalarial treatment compliance

#### Parenteral treatment

Across the full study period, most of the children were treated with an injectable antimalarial (DRC 83.7%, Nigeria 93.6% and Uganda 94.8%) without significant differences between community referrals and direct RHF attendances (Table 2). In 86.8% (6,153/7,088) of these cases, injectable artesunate was administered. During the post-RAS period, administration of parenteral antimalarials was higher (94.1%, artesunate 87.8%) than in the pre-RAS period (75.2%, artesunate 50.0%; Table 2, Figure 2). In DRC, the use of intravenous quinine was still common (18.3% among all children) though it was gradually replaced by artesunate throughout the study period (34.6% pre-RAS, 10.3% post-RAS). Before the roll-out of RAS, quinine use was relatively more common in children directly attending a RHF in DRC compared to community referrals (p < 0.001).

**Table 2:** Administration of antimalarial treatment for severe malaria, by country and enrolment level.

|  | Pooled (3 countries) |  |  | DRC |  |  | Nigeria |  |  | Uganda |  |  |
| --- | --- | --- | --- | --- | --- | --- | --- | --- | --- | --- | --- | --- |
|  | Community enrolments | RHF enrolments | P value (Chi2) | Community enrolments | RHF enrolments | P value (Chi2) | Community enrolments | RHF enrolments | P value (Chi2) | Community enrolments | RHF enrolments | P value (Chi2) |
|  | N = 2,381<br>n (%) | N = 5,602<br>n (%) |  | N = 1,939<br>n (%) | N = 2,269<br>n (%) |  | N = 217<br>n (%) | N = 834<br>n (%) |  | N = 225<br>n (%) | N = 2,449<br>n (%) |  |
| **Administration of at least one dose of an inj. antimalarial<sup>1</sup>** | 2030 (85.3) | 5058 (90.3) | <0.001 | 1613 (83.2) | 1908 (84.1) | 0.430 | 208 (95.9) | 776 (93.1) | 0.132 | 209 (92.9) | 2374 (95.0) | 0.171 |
| Artesunate | 1782 (74.8) | 4371 (78.0) | 0.002 | 1379 (71.1) | 1349 (59.5) | <0.001 | 202 (93.1) | 736 (88.3) | 0.040 | 201 (89.3) | 2286 (91.5) | 0.275 |
| Artemether | 17 (0.7) | 78 (1.4) | 0.011 | 11 (0.6) | 34 (1.5) | 0.003 | 6 (2.8) | 43 (5.2) | 0.137 | 0 (0.0) | 1 (0.0) | 0.764 |
| Quinine | 243 (10.2) | 646 (11.5) | 0.085 | 229 (11.8) | 539 (23.8) | <0.001 | 4 (1.8) | 8 (1.0) | 0.275 | 10 (4.4) | 99 (4.0) | 0.723 |
| **In-hospital administration of at least one dose of each an inj. and an oral antimalarial<sup>1</sup>** | 1374 (57.7) | 2665 (47.6) | <0.001 | 1300 (67.0) | 1499 (66.1) | 0.502 | 0 (0.0) | 28 (3.4) | 0.006 | 74 (32.9) | 1138 (45.5) | <0.001 |
| Artemether-lumefantrine<sup>2</sup> | 165 (6.9) | 1255 (22.4) | <0.001 | 91 (4.7) | 91 (4.0) | 0.278 | 0 (0.0) | 27 (3.2) | 0.007 | 74 (32.9) | 1137 (45.5) | <0.001 |
| Artesunate-amodiaquine<sup>2</sup> | 1010 (42.4) | 934 (16.6) | <0.001 | 1010 (52.1) | 930 (41.0) | <0.001 | 0 (0.0) | 1 (0.1) | 0.610 | 0 (0.0) | 0 (0.0) | NA |
| Quinine | 206 (8.7) | 495 (8.8) | 0.790 | 206 (10.6) | 494 (21.8) | <0.001 | 0 (0.0) | 0 (0.0) | NA | 0 (0.0) | 1 (0.0) | 0.764 |
| **Administration of ACT only** | 96 (4.0) | 126 (2.3) | <0.001 | 90 (4.6) | 57 (2.5) | <0.001 | 0 (0.0) | 4 (0.5) | 0.307 | 6 (2.7) | 65 (2.6) | 0.953 |

Number and column % of children receiving injectable and oral antimalarial treatment, pooled, by country and enrolment location
<sup>1</sup> More than one type of antimalarial may have been administered
<sup>2</sup> Compliant treatment

**Figure 2:**
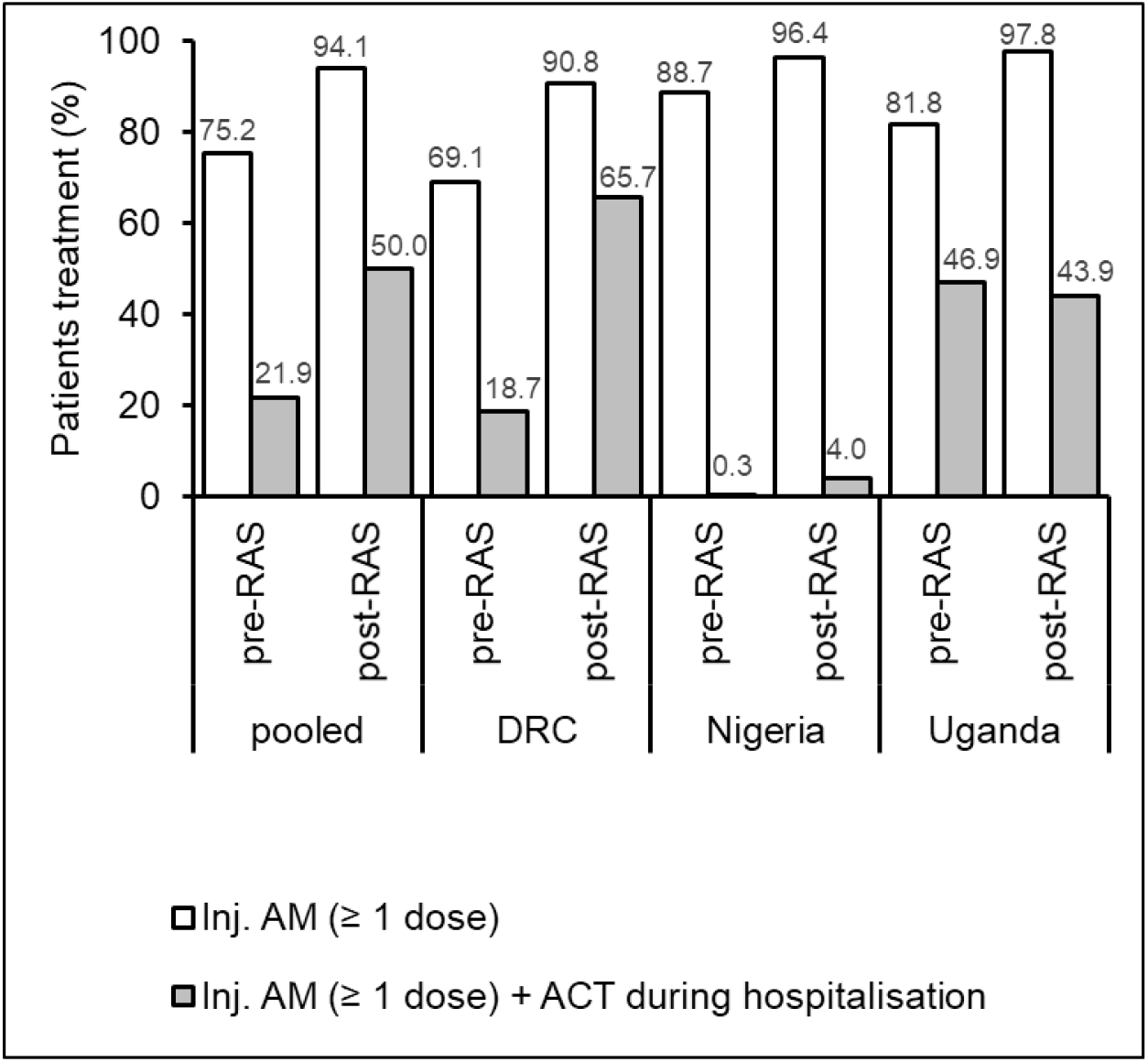
Antimalarial treatment compliance of children diagnosed with severe malaria before and after the implementation of RAS, pooled and by country (%). Proportion of children administered at least one dose of an injectable antimalarial (AM; artesunate, artemether or quinine; white bars), and administered each at least one dose of an injectable antimalarial and an in-hospital ACT of either ALU or ASAQ (compliant treatment; grey bars) before and after the roll-out of RAS.

#### ACT following parenteral treatment

Over the full study period, 50.6% of the children received both an injectable and an oral antimalarial while hospitalised: 42.0% were administered an ACT (17.8% ALU, 24.3% ASAQ), 8.8% received oral quinine, and 12 (0.15%) received both (Table 2). Large differences in oral follow-on treatment were observed between countries: only 2.7% of the children in Nigeria, 44.5% in Uganda and 50.3% in DRC were administered an ACT during admission. The levels of compliant treatment also varied considerably among included RHFs (21.6% - 68.0% in DRC, 0% - 7.5% in Nigeria and 19.4% - 88.7% in Uganda). ACT consisted mostly of ALU in Uganda and Nigeria (100% and 96.4% of all follow-on ACTs, respectively), children in DRC commonly received ASAQ (91.6%).

The pooled proportion of children receiving both, an injectable antimalarial and an ACT, increased significantly between the pre-RAS (21.9%) and the post-RAS period (50.0%, p < 0.001; Figure 2). This overall increase was mainly attributed to an increase of 47.0 and 3.7 percentage points (both p < 0.001) observed in DRC and Nigeria, respectively, as there was no difference between these two study periods in Uganda (46.9% vs. 43.9%). Children attending a community-based provider before referral to a RHF in Uganda and Nigeria were less likely to receive compliant treatment compared to direct RHF attendances (32.9% vs. 45.5%, p < 0.001 and 0% vs. 3.4%, p = 0.006, respectively; Table 2).

Follow-on treatment after injectable quinine consisted commonly of oral quinine in DRC (81.6% (627/768) received oral quinine vs. 6.3% who received ACT after quinine injection).

#### Predictors of compliant antimalarial treatment

Results from multivariable logistic regressions are presented in Table 3. While in Uganda and Nigeria, children of all ages were equally likely to receive compliant in-hospital treatment consisting of an injectable antimalarial followed by ACT, in DRC, the odds were higher for children below 3 years of age (aOR = 1.47, 95% CI 1.20-1.78 for children of 1 to 2 years relative to older children). Children in Uganda and Nigeria were equally likely to receive compliant treatment throughout the entire study period, whereas the odds were higher for children in DRC admitted during the post-RAS period (OR=6.19, 95% CI 4.81-7.95). While there was no difference in the odds of receiving compliant treatment between community referrals and direct RHF attendances in Uganda and DRC, referred children who received pre-referral treatment with RAS were more likely to receive an injectable followed by an ACT in DRC (aOR = 2.19, 95% CI 1.60-2.99) as compared to community referrals that were not treated with RAS. In Uganda, in contrast to the situation in DRC, children who received RAS were less likely to be administered compliant medication (aOR = 0.43, 95% CI 0.19-0.96). The low numbers of community referrals as well as low levels of compliant treatment observed in Nigeria did not allow to compute estimates for these indicators. Irrespective of malaria test result, all patients with a diagnosis of severe malaria at admission were equally likely to receive injectable treatment followed by an ACT. Other predictors for compliant treatment included costs incurred to caregivers: both in Uganda and DRC, payable hospitalisation was positively correlated with receiving injectable treatment and an ACT (aOR = 4.65, 95% CI 3.37-6.43 and aOR = 1.21, 95% CI 1.02-1.42 for Uganda and DRC, respectively), while the odds were lower if caregivers had to pay for drugs (aOR = 0.25, 95% CI 0.20-0.33 and aOR = 0.81, 95% CI 0.78-0.97 for Uganda and DRC, respectively). Again, the number of events in Nigeria did not permit estimating these predictors. The odds of receiving compliant in-hospital treatment varied substantially among the study areas within the countries; in the DRC, being enrolled in Kingandu health zone was negatively associated with provision of compliant in-hospital medication compared to Ipamu and Kenge health zones (aOR = 0.67, 95% CI 0.54 -0.84). Treatment compliance was much better in Song LGA in Nigeria (aOR = 10.3, 95% CI 1.33 -79.6) and in both Oyam (aOR = 5.34, 95% CI 4.17-6.87) and Apac districts (aOR = 5.80, 95% CI 4.17-8.08) in Uganda.

**Table 3:**
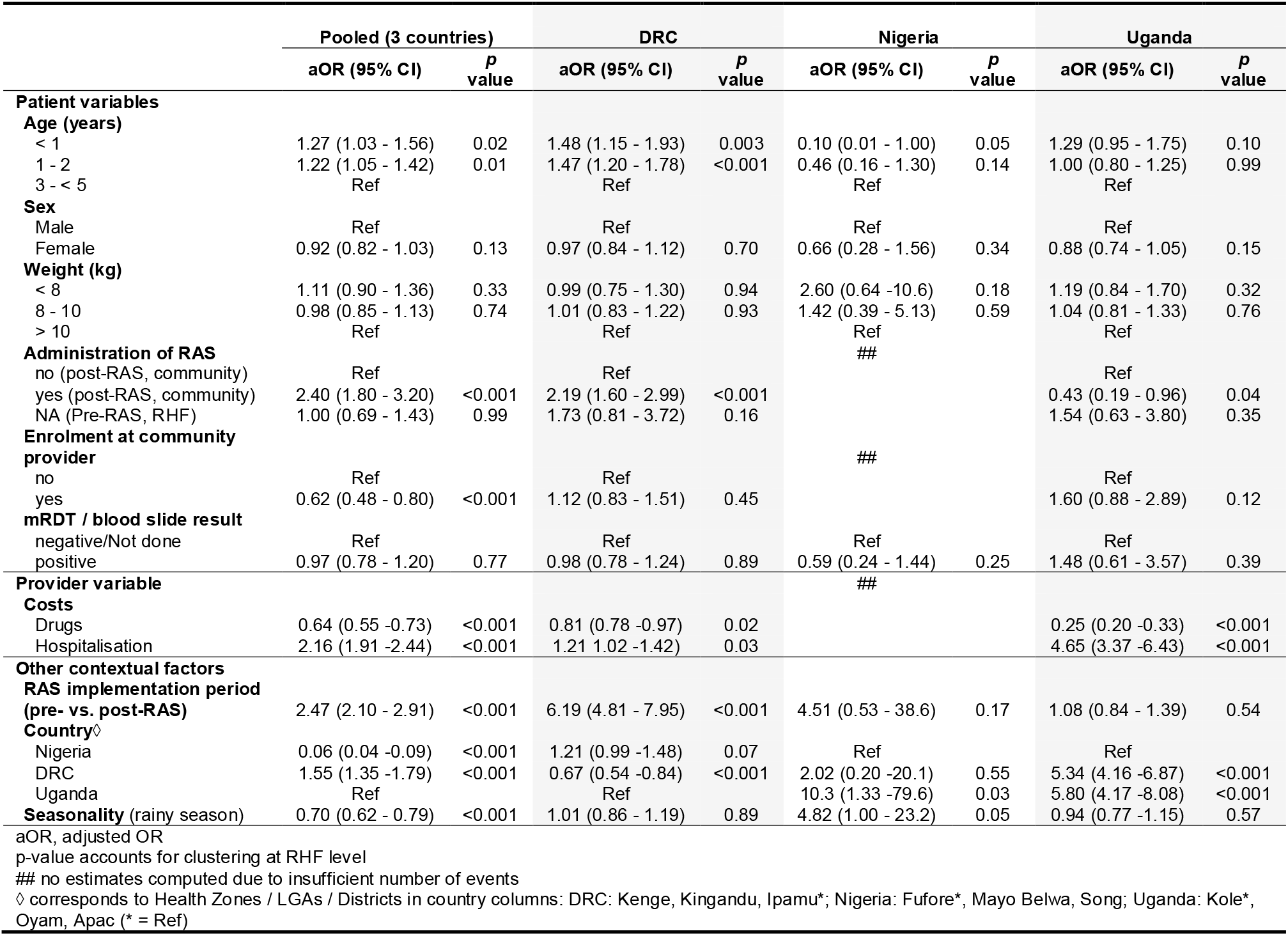
Patient, provider, and facility correlates with antimalarial treatment compliance according to the WHO malaria treatment guidelines (logistic regression).

#### In-hospital versus home ACT treatment

Study countries differed considerably in the way of providing ACT following parenteral treatment (Table S3). While in DRC, ACT was usually started as an in-hospital therapy following completion of injectable treatment (78.7%, 1,314/1,669) and completed in 49.3% (822/1,669) of the cases while the patient was still hospitalised, only 1.7% (7/421) in Nigeria and 45.7% (617/1,349) in Uganda received any ACTs as in-patients. 12.8% completed ACT treatment before discharge in Uganda, not a single child did so in Nigeria. Issuing prescriptions to buy ACTs at discharge was much more common in Nigeria (54.4% of admitted children) and Uganda (53.0%). In Uganda, relatively more children referred to a RHF by a community-based provider received a prescription (68.9%) instead of in-hospital ACT treatment (30.3%) compared to children directly attending a RHF (47.4% started ACT therapy while hospitalised, 51.3% received a prescription at discharge).

#### Health worker compliance of antimalarial treatment administration and treatment prescription

Among the post-RAS subsample with detailed dosage information (N=3,449), the vast majority treated with an injectable antimalarial received at least three doses (3,220/3,337 (96.5%); Table S4). In DRC, 76.2% of children were subsequently treated with an ACT during admission (Figure 4); the percentage of compliant treatment prescription (i.e. administration or prescription or dispensing of an ACT after injectable treatment) remained basically unchanged (76.9%) due to the low levels of post-discharge ACT prescription. While in Nigeria, the level of compliant in-hospital treatment administration was very low (1.2%), compliant antimalarial prescription was increased but remained low with only 45.6% in the post-RAS period. And finally, including ACT prescriptions in the estimate for compliant treatment more than doubled the percentage for Uganda (44.7% vs. 97.5%). Compliance of in-hospital treatment administration to children referred from the community was lower in Uganda as compared to direct RHF attendances (29.8% vs. 46.7%, p < 0.001; Table S4). Since community referrals are more likely to receive a prescription, this difference vanished for compliant treatment prescription (99.2% vs. 98.2%).

**Figure 3:**
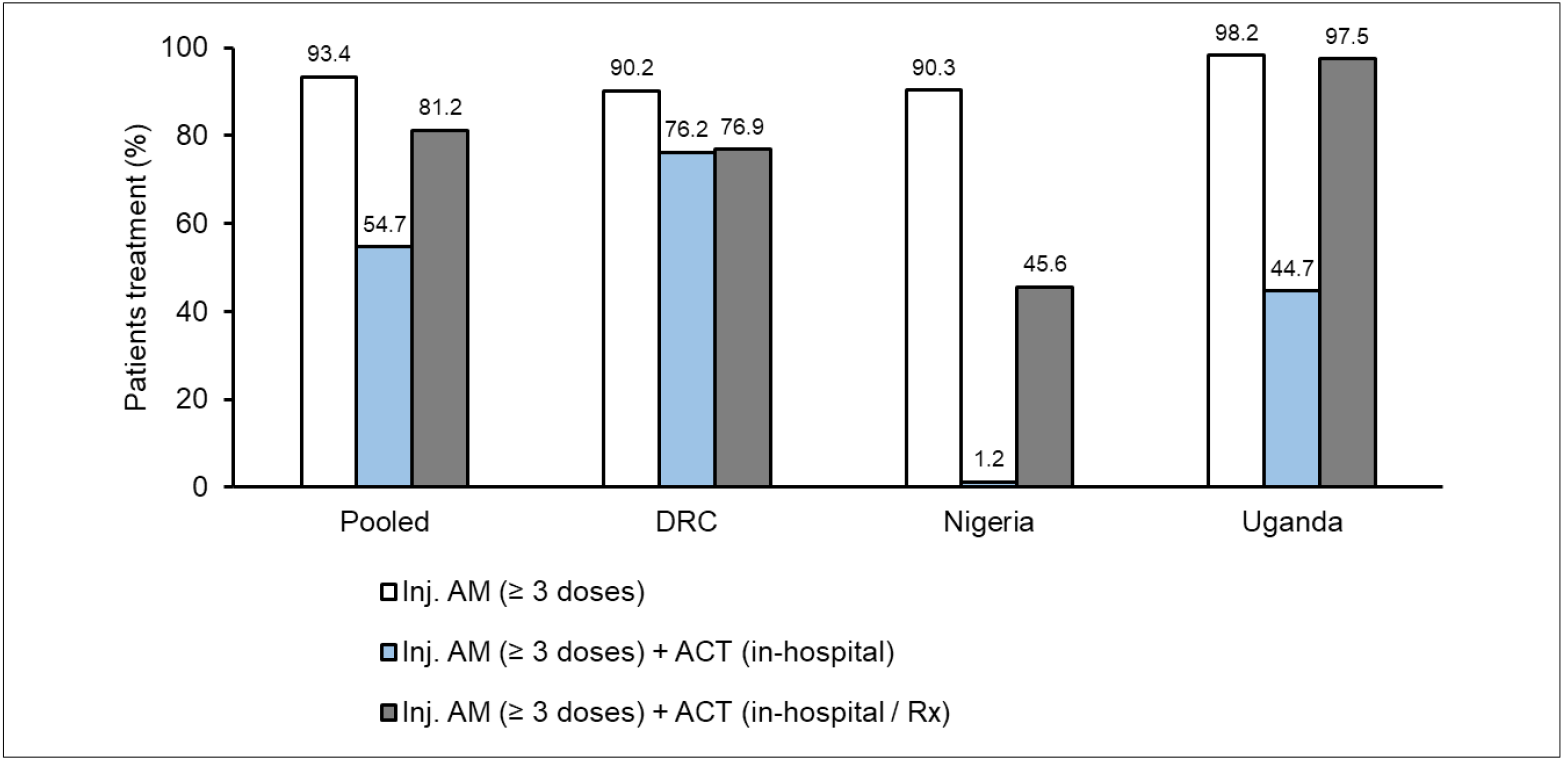
Treatment compliance for children diagnosed with severe malaria after the implementation of RAS, pooled and by country (%). Proportion of children administered at least 3 doses of an injectable antimalarial (AM; artesunate, artemether or quinine; white bars), at least 3 doses of an injectable antimalarial and in-hospital follow-on ACT (compliant treatment administration; blue bars) or in-hospital administered / at discharge prescribed / dispensed follow-on ACT (compliant treatment prescription; dark grey bars). Data collection period: Uganda and DRC: April 2019 - July 2020, Nigeria: May 2019 - July 2020. Rx: prescription.

**Figure 4:**
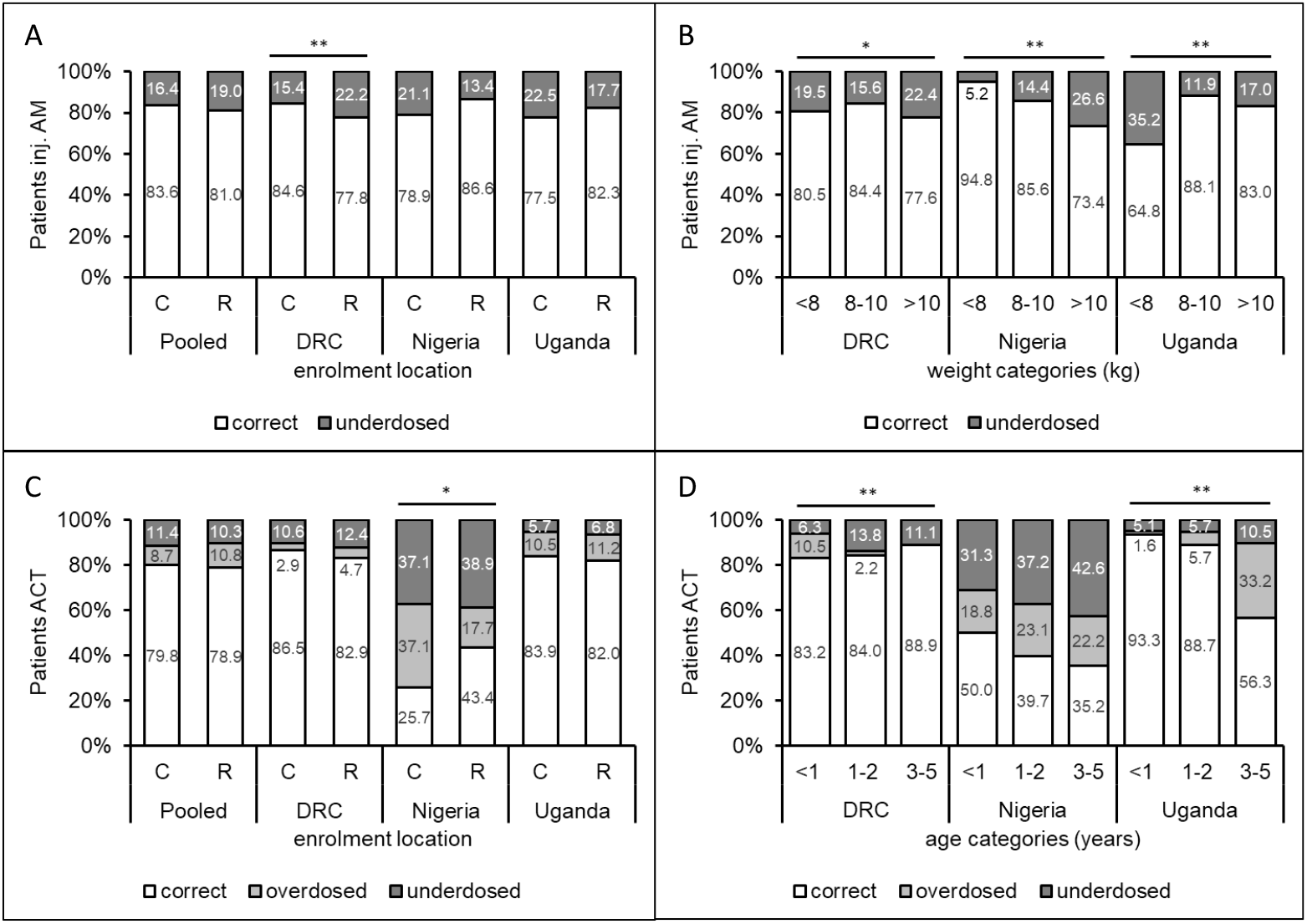
Prescription of injectable antimalarials and ACTs in compliant total doses (%). Administration of inj. antimalarial (A, B) and prescription of ACTs (C, D) in compliant doses by enrolment location (A, C) and weight / age category (B, D). Compliant dosing was defined as administration of the equivalent of min. 3 doses of an injectable antimalarial (artesunate, artemether or quinine) at the WHO-recommended dose (correct minimal total dose). ACT dosing was considered correct if ALU / ASAQ was prescribed / dispensed for 3 days at the recommended dose (see Supplementary materials S5). Inj. AM dosing data was missing for 2.2% in DRC, 30.8% in Nigeria and 30.3% in Uganda. Complete ACT dosing information was missing for 67.9% in DRC, 25.6% in Nigeria and 13.1% in Uganda. C, community enrolments; R, RHF enrolments; * p < 0.05, ** p ≤ 0.001.

While complete dosage information for in-hospital ACT treatment was not available, the majority of ACT prescriptions / dispensing at discharge were for a three-day treatment course (1,664/1,730, 96.2%; Table S4).

### Dosing compliance

Compliance with the WHO dosing recommendations for parenteral antimalarials was estimated by the total amount of drug administered relative to the child’s body weight (missing = 570, 16.5%). Dosing compliance for injectable artesunate was high in all three countries (82.2% in Uganda, 84.3% in DRC and 84.9% in Nigeria; Table S5). Levels of correct dosing were lower in RHF enrolments in DRC (81.4%) as compared to community enrolments (87.0%, p = 0.003). While only relatively few children in this study period received quinine, dosing of this drug was often not compliant with the WHO guidelines: 84.7% (61/72), 100% (2/2) and 66.7% (6/9) of children with quinine dosing data available in DRC, Nigeria and Uganda, respectively, did not receive the correct minimal total dose. While in Nigeria, there seems to be a tendency to under-dose injectables in children with a higher weight, the opposite was found in Uganda, where children of lesser weight more often received an insufficient total dose (Figure 4). Most children under-dosed still received at least three doses (Table S5). It is worth mentioning that during the respective study period, 83 (2.4%) children deceased during hospitalisation, 85.7% of them within three days of admission. Excluding these children from the dosing compliance analysis for injectable treatment did not substantially affect these results (data not shown).

While prescription / dispensing of 3-day ACT treatment courses were common (96.2%; Table S4), the proportion of correct total dosing of ACTs (according to the child’s age, Table S1) ranged from 39.2% in Nigeria, 82.2% in Uganda to 84.8% in DRC. In Nigeria, a higher proportion of RHF enrolments was correctly dosed (43.4%) as compared to community enrolments (25.7%, p = 0.035; Figure 4, Table S5); furthermore, children below one year of age were more likely to receive a correct total ACT dose (50.0%), compared to children between 1 and 2 years old (39.7%) and 3 to 5 years old (35.2%). In DRC, correct ACT dosing remained high for all ages and was highest for the oldest age category (88.9%). In Uganda, children below 1 year of age received a correct ACT prescription / dispensing in 93.3% of cases, but correct dosing declined for older age categories (88.7% for children between 1 and 2 years, 56.3% for children between 3 and 5 years).

## Discussion

The CARAMAL study aimed to understand healthcare seeking, provision of antimalarial treatment and health outcomes of children suffering from severe malaria in the context of the integration of pre-referral RAS into the continuum of severe malaria care. Adequate post-referral management of severe malaria at the designated health facilities is critical to ensure complete patient cure and avoid death and persisting disability.

A number of studies have previously addressed health worker treatment compliance for both uncomplicated (19-31) and severe malaria (7, 8, 10, 32-34). However, the CARAMAL project was special for several reasons: Firstly, it is the first systematic evaluation of post-referral treatment of severe malaria following the roll-out of pre-referral RAS. Secondly, the study was very large, with clinical observation data from 7,983 hospitalisations for severe malaria at 25 referral health facilities (RHFs) in three highly malaria-endemic African countries. Thirdly, recruitment was broad at both, community and RHF level, and a visit at home of the patients 28 days after treatment initiation allowed an assessment of treatment outcome, as well as the triangulation of multiple findings.

Completion of severe malaria treatment with an ACT is a central component of the WHO recommendation. Our results show that compliance with this recommendation is rather low: only 1.2% of children undergoing treatment at a RHF after the roll-out of RAS in Nigeria received three doses of injectable antimalarial followed by an ACT during hospitalisation. This percentage was also unsatisfactory in Uganda with 44.7% and reached 76.2% in DRC.

Published results for this indicator vary greatly ranging between 4.8% in Uganda (8) and 43.4% in Nigeria (33) of patients having received in-hospital injectable antimalarials followed by a co-prescription of oral ACT, though these reports do not clearly specify whether ACTs were prescribed for in-hospital administration or at discharge. In our study, we found that all three countries adopted different methods of ACT provision: either, ACTs were directly administered at the RHF or patients were discharged with a prescription for home treatment, or a variation of these.

Provision of ACT as prescription or, as in Nigeria, discharging children without ACT altogether (43.9%), in addition to the low dosing compliance for these drugs, raises major concerns about the effectiveness of the treatment that these children receive during their episode of severe malaria. Artemisinin monotherapy and incomplete ACT treatment bears a high risk not only for incomplete parasite clearance and recrudescence of the infection, but also for the development or selection of artemisinin resistant parasites. The selection of *P. falciparum* harbouring artemisinin resistant parasites has been found in the context of the CARAMAL project in Uganda (Awor et al., manuscript in preparation).

If treatment is provided as prescription the patient’s or caregiver’s adherence to the correct dose is crucial to ensure effective antimalarial treatment. Studies on patient and caregiver adherence to antimalarial treatment guidelines found large variations among different countries, ranging from <50% to 100% (35-38). For Uganda, a range between 65.8% and 99.2% was reported. One study conducting household interviews in Nigeria presented an adherence of 93.3% (39). Siddiqui et al. (31) reported 62% patient adherence to ACT treatment for uncomplicated malaria in DRC. Adherence may also be influenced by whether ACTs are delivered by the public or the private sector as well as by caregiver income (37). However, all authors acknowledged the weak evidence and challenges and weaknesses of adherence measures (e.g. self-reporting, pill counts, bio-assays, etc.). Certainly, more research is required in this direction.

One reason for discharging a child before the start of ACT therapy could lie in a limited bed capacity of the RHFs, especially during rainy seasons or disease outbreaks. Though we did not analyse this further, COVID-19 had an impact on the running of these facilities in both Nigeria and Uganda (e.g. restrictions in Nigeria and Uganda impacting the movement of persons and the supply chain between March and July 2020). ACT stock-outs may also have led to an increase of prescriptions (CARAMAL project, data not yet analysed). Further, socioeconomic factors may have influenced differently hospitalisation duration and treatment patterns among community and RHF enrolments. Such factors may account for differential methods for ACT provision in Uganda, where referral cases were more likely to receive a prescription rather than in-hospital ACT treatment compared to children directly attending a RHF. Early hospital discharge and provision of ACT as prescription may also be because inpatient medical care is no longer absolutely required (e.g. the child is able to swallow and further treatment may be provided by the caregiver). Discharging therefore relieves the burden on both the RHF (beds) and the family (costs) – but at the potential expense of ACT adherence.

The relatively high rate of treatment compliance observed in DRC during the post-RAS period may be a result of a number of supportive interventions implemented to facilitate the roll-out and uptake of RAS (Lengeler, Burri et al. and Lambiris et al., manuscripts submitted). In particular, this included distributing injectable artesunate to referral facilities in DRC (27,000 artesunate vials in 2019) to limit use of the inferior quinine (40-42). This increased availability of artesunate led to the gradual replacement of quinine in DRC (34.6% treated with IV quinine pre-RAS vs. 10.3% post-RAS). Safe and effective antimalarial treatment for severe malaria will need a sustained procurement, distribution and use of artesunate beyond the end of the project.

Administration of pre-referral RAS did have a positive impact on whether a child receives compliant treatment composed of injectable antimalarials and ACT in DRC. The opposite was the case for Uganda, where RAS was negatively correlated with treatment compliance. This finding raises important concerns, namely that the full course of antimalarial treatment for severe disease may no longer be considered necessary after a dose of RAS and a possible rapid improvement of the child’s health condition. This aspect should be emphasized in health worker trainings. Our finding that the likelihood of receiving compliant treatment increased if caregivers had to pay a hospitalisation fee seems to imply that good quality of care has its cost.

In line with previous studies (7, 8, 10, 34), our data showed that despite the variations in treatment compliance, the majority of children undergoing treatment for severe malaria at a RHF received an injectable antimalarial (88.8%), which was mostly artesunate (77.1%). In all three countries, children referred from the community were as likely to receive parenteral treatment as children directly attending a RHF suggesting equal management of these children at the RHF level.

Weight-based dosing compliance for injectable artesunate was 82.2% in Uganda, 84.3% in DRC and 84.9% in Nigeria. Two earlier reports from Uganda showed that the artesunate dose and dosing scheme followed the recommendations in 70.4% of patients (7), and 65.6% of weighted children <20 kg were prescribed the correct dose (34). Health worker training and supervision promoted by CARAMAL may have contributed to the comparably high dosing compliance in our study. On the other hand, dosing compliance was poor for parenteral quinine. It is worth mentioning, that both DRC and Uganda follow quinine treatment guidelines that are not aligned with the WHO dose recommendation for the loading dose (S1 Table). However, based on country-specific dosing recommendations, still 51.4% (37/72, data not shown) of children in DRC did not receive the correct minimal total dose (in 89.2% of these cases this was due to not receiving a minimum of three doses; data not shown). Overall, most of the children who received a too low total dose of injectable treatment were administered three doses, pointing to a problem with calculating the correct individual dose. Cross-sectional healthcare provider surveys conducted in the frame of the CARAMAL study found most RHFs to be equipped with a functional weighing scale (CARAMAL project, unpublished data). However, not all children may have been weighed but weight approximations applied, e.g. the Advanced Paediatric Life Support (APLS) age-for-weight estimation (Weight 1 - 5 = (Age x 2) + 8). Issues with correct age may arise in situations where birth registration and date-age calculations are inaccurate, leading to low levels of age-based dosing compliance like observed in Nigeria (39.2%).

While our study lacks information on patient adherence with prescribed ACT, the prospective recording of case management during hospitalisation provided more accurate treatment information than other studies using a retrospective design. The multi-country study allowed us to include an unprecedented large sample of severely ill children from very distinct contexts (in terms of disease burden, health system, access to healthcare, etc.) while at the same time investigating the effect of introducing pre-referral RAS.

At the same time, the different contexts in each country (incl. the health system and research set-up) led to slight differences in the detail of data collected, impacting the depth in which certain findings could be analysed across countries. Intensive training of data collection staff and standardized record forms were implemented to minimise observer bias and differences between countries. We cannot rule out the possibility of a Hawthorne effect due to the study staff’s presence, potentially leading to an overestimation of the true quality of care.

This study was limited to evaluating the treatment of severe malaria in patients included based on the local clinicians’ diagnosis of “severe malaria”. This diagnosis was not independently confirmed. In addition, a large proportion of children diagnosed with severe malaria in the RHFs (health centre IV) in Uganda received outpatient antimalarial treatment instead of being admitted. These patients as well as those who were treated for severe malaria by other health care providers were not included in this study. We cannot rule out that antimalarial treatment and compliance to the WHO recommendations in these facilities may differ from the treatment observed in this study.

## Conclusion

The quality of antimalarial treatment provided at the referral facility must be improved to meet the WHO treatment recommendations. While parenteral treatment was administered correctly and reliably, we found that the mandatory provision of ACTs to complete treatment was often not followed or left to the discretion of the caregiver for home treatment. Pre-referral RAS for children in hard-to-reach locations can only be an effective component of the continuum of care for severe malaria if post-referral treatment is adequate.

## Data Availability

All data produced in the present study are available upon reasonable request to the authors.

## Acknowledgements

The study team would like to sincerely thank all the children and their caregivers who agreed to participate in this study; the health workers and local and national health authorities who provided their support; our study teams of the Kinshasa School of Public Health (DRC), Akena Associates Ltd. (Nigeria), and the Makerere University School of Public Health (Uganda); and the colleagues of the local CHAI and UNICEF offices.

## Supporting information

**Table S1:** Treatment and dose recommendation for severe malaria in children < 5 years.

**Table S2:** Summary characteristics of surveyed patients and exposure variables (subsample, post-RAS only).

**Table S3:** Provision of in-hospital vs. post-discharge ACT medication.

**Table S4:** Antimalarial treatment compliance: number of doses and total doses of injectable antimalarials administered and ACTs prescribed (subsample, post-RAS only)

**Table S5:** Antimalarial dosing compliance: total doses of injectable antimalarials administered and ACTs prescribed (subsample, post-RAS only).

**Supplementary materials S6:** Assessment of correct dosing of injectable antimalarials and ACTs.

## References

1. Mousa A, Al-Taiar A, Anstey NM, Badaut C, Barber BE, Bassat Q, et al. The impact of delayed treatment of uncomplicated P. falciparum malaria on progression to severe malaria: A systematic review and a pooled multicentre individual-patient meta-analysis. PLOS Medicine. 2020;17(10):e1003359.

2. WHO. Severe malaria. Trop Med Int Health. 2014;19 Suppl 1:7–131.

3. World Health Organization. Guidelines for the treatment of malaria. 3rd ed: World Health Organization; 2015.

4. Camponovo F, Bever CA, Galactionova K, Smith T, Penny MA. Incidence and admission rates for severe malaria and their impact on mortality in Africa. Malaria Journal. 2017;16(1):1.

5. Kadobera D, Sartorius B, Masanja H, Mathew A, Waiswa P. The effect of distance to formal health facility on childhood mortality in rural Tanzania, 2005-2007. Glob Health Action. 2012;5:1–9.

6. Schoeps A, Gabrysch S, Niamba L, Sie A, Becher H. The effect of distance to health-care facilities on childhood mortality in rural Burkina Faso. Am J Epidemiol. 2011;173(5):492–8.

7. Achan J, Tibenderana J, Kyabayinze D, Mawejje H, Mugizi R, Mpeka B, et al. Case Management of Severe Malaria - A Forgotten Practice: Experiences from Health Facilities in Uganda. PLoS ONE. 2011;6(3):e17053.

8. Ampadu HH. Prescribing patterns and compliance with World Health Organization recommendations for the management of severe malaria: a modified cohort event monitoring study in public health facilities in Ghana and Uganda. 2019:8.

9. Clarke-Deelder E, Shapira G, Samaha H, Fritsche GB, Fink G. Quality of care for children with severe disease in the Democratic Republic of the Congo. BMC Public Health. 2019;19(1):1608.

10. Shah MP, Briggs-Hagen M, Chinkhumba J, Bauleni A, Chalira A, Moyo D, et al. Adherence to national guidelines for the diagnosis and management of severe malaria: a nationwide, cross-sectional survey in Malawi, 2012. Malaria Journal. 2016;15(1):369.

11. Gomes MF, Faiz MA, Gyapong JO, Warsame M, Agbenyega T, Babiker A, et al. Pre-referral rectal artesunate to prevent death and disability in severe malaria: a placebo-controlled trial. Lancet. 2009;373(9663):557–66.

12. Medicines for Malaria Venture. Strides Shasun’s rectal artesunate product receives WHO prequalification 2018 [Available from: https://www.mmv.org/newsroom/news/strides-shasun-s-rectal-artesunate-product-receives-who-prequalification.

13. CARAMAL consortium. Rectal Artesunate Landscaping Assessment Report. 2018 [Available from: https://www.severemalaria.org/sites/mmv-smo/files/content/attachments/2019-02-20/RAS%20landscaping%20report_2018_updated.pdf.

14. von Seidlein L, Deen JL. Pre-referral rectal artesunate in severe malaria. Lancet. 2009;373(9663):522–3.

15. Brunner NC, Omoluabi E, Awor P, Okitawutshu J, Tshefu A, Signorell A, et al. Pre-referral rectal artesunate and referral completion among children with suspected severe malaria in the Democratic Republic of the Congo, Nigeria and Uganda 2021. medRxiv [Preprint]. medRxiv 2021.09.27.21264073; doi: https://doi.org/10.1101/2021.09.27.21264073 [posted October 15, 2021] Available from: https://www.medrxiv.org/content/10.1101/2021.09.27.21264073v2.full.pdf.

16. Hetzel MW, Okitawutshu J, Tshefu A, Omoluabi E, Awor P, Signorell A, et al. Effectiveness of rectal artesunate as pre-referral treatment for severe malaria in children <5 years of age 2021. medRxiv [Preprint]. medRxiv 2021.09.24.21263966; doi: https://doi.org/10.1101/2021.09.24.21263966 [posted September 27, 2021] Available from: https://www.medrxiv.org/content/10.1101/2021.09.24.21263966v1.full.pdf.

17. Idro R, Aketch S, Gwer S, Newton CR, Maitland K. Research priorities in the management of severe Plasmodium falciparum malaria in children. Ann Trop Med Parasitol. 2006;100(2):95–108.

18. World Health Organization. Guidelines for malaria. Geneva. 2021.

19. Amboko B, Stepniewska K, Macharia PM, Machini B, Bejon P, Snow RW, et al. Trends in health workers’ compliance with outpatient malaria case-management guidelines across malaria epidemiological zones in Kenya, 2010–2016. Malaria Journal. 2020;19(1):406.

20. Bamiselu OF. Adherence to malaria diagnosis and treatment guidelines among healthcare workers in Ogun State, Nigeria. 2016:10.

21. Bawate C. Factors affecting adherence to national malaria treatment guidelines in management of malaria among public healthcare workers in Kamuli District, Uganda. 2016:10.

22. Budimu A, Emidi B, Mkumbaye S, Kajeguka DC. Adherence, Awareness, Access, and Use of Standard Diagnosis and Treatment Guideline for Malaria Case Management among Healthcare Workers in Meatu, Tanzania. Journal of Tropical Medicine. 2020;2020:1–6.

23. Camara A, Moriarty LF, Guilavogui T, Diakité PS, Zoumanigui JS, Sidibé S, et al. Prescriber practices and patient adherence to artemisinin-based combination therapy for the treatment of uncomplicated malaria in Guinea, 2016. Malaria Journal. 2019;18(1):23.

24. Cohen JL, Leslie HH, Saran I, Fink G. Quality of clinical management of children diagnosed with malaria: A cross-sectional assessment in 9 sub-Saharan African countries between 2007–2018. PLOS Medicine. 2020;17(9):e1003254.

25. Ezenduka CC, Okonta MJ, Esimone CO. Adherence to treatment guidelines for uncomplicated malaria at two public health facilities in Nigeria; Implications for the ‘test and treat’ policy of malaria case management. Journal of Pharmaceutical Policy and Practice. 2014;7(1):15.

26. Kaula H. Cross-sectional study on the adherence to malaria guidelines in lakeshore facilities of Buyende and Kaliro districts, Uganda. 2018:9.

27. Masanja IM, Selemani M, Khatib RA, Amuri B, Kuepfer I, Kajungu D, et al. Correct dosing of artemether-lumefantrine for management of uncomplicated malaria in rural Tanzania: do facility and patient characteristics matter? Malaria Journal. 2013;12(1):446.

28. Namuyinga RJ, Mwandama D, Moyo D, Gumbo A, Troell P, Kobayashi M, et al. Health worker adherence to malaria treatment guidelines at outpatient health facilities in southern Malawi following implementation of universal access to diagnostic testing. Malaria Journal. 2017;16(1):40.

29. Amboko BI, Ayieko P, Ogero M, Julius T, Irimu G, English M. Malaria investigation and treatment of children admitted to county hospitals in western Kenya. Malaria Journal. 2016;15(1):506.

30. Selemani M, Masanja IM, Kajungu D, Amuri M, Njozi M, Khatib RA, et al. Health worker factors associated with prescribing of artemisinin combination therapy for uncomplicated malaria in rural Tanzania. Malaria Journal. 2013;12(1):334.

31. Siddiqui MR, Willis A, Bil K, Singh J, Mukomena Sompwe E, Ariti C. Adherence to Artemisinin Combination Therapy for the treatment of uncomplicated malaria in the Democratic Republic of the Congo. F1000Res. 2015;4:51.

32. Makumbe B, Tshuma C, Shambira G, Mungati M, Gombe NT, Bangure D, et al. Evaluation of severe malaria case management in Mazowe District, Zimbabwe, 2014. Pan African Medical Journal. 2017;27.

33. Ojo AA, Maxwell K, Oresanya O, Adaji J, Hamade P, Tibenderana JK, et al. Health systems readiness and quality of inpatient malaria case-management in Kano State, Nigeria. Malaria Journal. 2020;19(1):384.

34. Zurovac D, Machini B, Kiptui R, Memusi D, Amboko B, Kigen S, et al. Monitoring health systems readiness and inpatient malaria case-management at Kenyan county hospitals. Malar J. 2018;17(1):213.

35. Banek K, Webb EL, Smith SJ, Chandramohan D, Staedke SG. Adherence to treatment with artemether–lumefantrine or amodiaquine–artesunate for uncomplicated malaria in children in Sierra Leone: a randomized trial. Malaria Journal. 2018;17(1):222.

36. Bruxvoort K, Goodman C, Kachur SP, Schellenberg D. How Patients Take Malaria Treatment: A Systematic Review of the Literature on Adherence to Antimalarial Drugs. PLoS ONE. 2014;9(1):e84555.

37. Yakasai AM, Hamza M, Dalhat MM, Bello M, Gadanya MA, Yaqub ZM, et al. Adherence to Artemisinin-Based Combination Therapy for the Treatment of Uncomplicated Malaria: A Systematic Review and Meta-Analysis. Journal of Tropical Medicine. 11.

38. Banek K, Lalani M, Staedke SG, Chandramohan D. Adherence to artemisinin-based combination therapy for the treatment of malaria: a systematic review of the evidence. 2014:14.

39. Ajayi IO, Browne EN, Bateganya F, Yar D, Happi C, Falade CO, et al. Effectiveness of artemisinin-based combination therapy used in the context of home management of malaria: a report from three study sites in sub-Saharan Africa. Malar J. 2008;7:190.

40. Conroy AL, Opoka RO, Bangirana P, Namazzi R, Okullo AE, Georgieff MK, et al. Parenteral artemisinins are associated with reduced mortality and neurologic deficits and improved long-term behavioral outcomes in children with severe malaria. BMC Med. 2021;19(1):168.

41. Dondorp A, Nosten F, Stepniewska K, Day N, White N, South East Asian Quinine Artesunate Malaria Trial g. Artesunate versus quinine for treatment of severe falciparum malaria: a randomised trial. Lancet. 2005;366(9487):717–25.

42. Dondorp AM, Fanello CI, Hendriksen IC, Gomes E, Seni A, Chhaganlal KD, et al. Artesunate versus quinine in the treatment of severe falciparum malaria in African children (AQUAMAT): an open-label, randomised trial. Lancet. 2010;376(9753):1647–57.

