## Supporting information for "Health worker compliance with severe malaria treatment guidelines in the context of implementing pre-referral rectal artesunate: an operational study in three high burden countries"

**Table S1:** Treatment and dose recommendation for severe malaria in children < 5 years.

| | First-line parenteral treatment | Treatment schedule | Dosage | Second-line parenteral treatment | Treatment schedule | Dosage |
|---|---|---|---|---|---|---|
| **WHO** | AS | At 0 hours<br>At 12 hours<br>At 24 hours | Child less than 20 kg: 3 mg / kg<br>Adults and child > 20 kg: 2.4 mg / kg | Artemether (if parenteral AS is not available) | At 0 hours | 3.2 mg / kg |
| | | | | | At 24 hours | 1.6 mg / kg |
| | | | | | At 48 hours | 1.6 mg / kg |
| | | Then once a day until patient is able to tolerate oral medication, then give a full course of oral ACT | | | Then once a day until patient is able to tolerate oral medication, then give a full course of oral ACT | |
| | | | | Quinine (use artemether in preference to quinine) | At 0 hours | 20 mg / kg |
| | | | | | At 8 hours | 10 mg / kg |
| | | | | | Then 10 mg / kg every 8 hours until patient is able to tolerate oral medication, then give a full course of oral ACT | |
| **DRC** | AS | Same like WHO | | Quinine (if AS is contra-indicated) | At 0 hours | 10 mg / kg |
| | | | | | At 12 hours | 10 mg / kg |
| | | | | | Then 10 mg / kg every 12 hours until patient is able to tolerate oral medication, then give a full course of oral ACT or oral quinine + clindamycin to complete 7 days of quinine therapy | |
| **Nigeria** | AS | Same like WHO | | Artemether(if parenteral AS is not available) | Same like WHO | |
| | | | | Quinine(if parenteral AS is not available) | Same like WHO | |
| **Uganda** | AS | Same like WHO | | Artemether | Same like WHO | |
| | | | | Quinine | At 0 hours | 10 mg / kg |
| | | | | | At 8 hours | 10 mg / kg |
| | | | | | Then every 8 hours until when patient is able to take oral medication | |

| | First-line ACT | Treatment schedule | Dosage | Alternative first-line ACT |
|---|---|---|---|---|
| **WHO** | ALU | 3 days | 5 to < 15 kg : 1 x 20/120 mg ALU twice daily | ASMQ, DHA-PQ, AS-SP |
| | | | 15 to < 25 kg: 2 x 20/120 mg ALU twice daily | |
| | ASAQ | 3 days | 4.5 to < 9 kg: 1 x 25/67.5 mg ASAQ once daily | |
| | | | 9 to < 18 kg: 1 x 50/135 mg ASAQ once daily | |
| **DRC** | ASAQ | 3 days | 4.5 to < 9 kg / 2 to 11 months: 1 x 25/67.5 mg ASAQ once daily | none |
| | | | 9 to < 18 kg / 1 to 5 years: 1 x 50/135 mg ASAQ once daily | |

| | | | | |
|---|---|---|---|---|
| | ALU | Same like WHO | | |
| Nigeri | ALU | Same like WHO | | none |
| | ASAQ | Same like WHO | | |
| Uganda | ALU | 3 days | 5 to 14 kg / 4 months to 3 years: 1 x 20/120 mg ALU twice daily | DHA-PQ |
| | | | 15 to 24 kg / 3 to 7 years: 2 x 20/120 mg ALU twice daily | |
| | ASAQ | 3 days | 5 to 11 months: 25/76 mg ASAQ once daily | none |
| | | | 1 to 6 years: 50/153 mg ASAQ once daily | |

Guidelines for the treatment of malaria issued by the WHO (1) and the study countries DRC (2), Nigeria (3) and Uganda (4). ACT, artemisinin-based combination therapy; ALU, artemether-lumefantrine; ASAQ, artesunate-amodiaquine; DHA-PQ, dihydroartemisinin-piperaquin, AS-SP, artesunate+sulfadoxine-pyrimethamine

**Table S2:** Summary characteristics of surveyed patients and exposure variables (subsample, post-RAS only).

| | Pooled (3 countries) | | DRC | | Nigeria | | Uganda | |
|---|---|---|---|---|---|---|---|---|
| | Community enrolments N = 1,095 n (%) | RHF enrolments N = 2,354 n (%) | Community enrolments N = 850 n (%) | RHF enrolments N = 823 n (%) | Community enrolments N = 113 n (%) | RHF enrolments N = 308 n (%) | Community enrolments N = 132 n (%) | RHF enrolments N = 1,223 n (%) |
| **Age (years)** | | | | | | | | |
| < 1 | 237 (21.6) | 496 (21.1) | 199 (23.4) | 175 (21.3) | 13 (11.5) | 42 (13.6) | 25 (18.9) | 279 (22.8) |
| 1 - 2 | 576 (52.6) | 1247 (53.0) | 427 (50.2) | 412 (50.1) | 70 (62.0) | 181 (58.8) | 79 (59.9) | 654 (53.5) |
| 3 - < 5 | 282 (25.8) | 611 (26.0) | 224 (26.4) | 236 (28.7) | 30 (26.6) | 85 (27.6) | 28 (21.2) | 290 (23.7) |
| **Sex** | | | | | | | | |
| Female | 514 (46.9) | 1058 (44.9) | 415 (48.8) | 375 (45.6) | 77 (31.9) | 140 (45.5) | 63 (47.7) | 543 (44.4) |
| **Weight (kg)** | | | | | | | | |
| < 8 | 178 (17.9) | 331 (17.6) | 155 (18.4) | 113 (14.1) | 12 (16.7) | 67 (30.6) | 11 (13.6) | 151 (17.5) |
| 8 - 10 | 467 (46.9) | 816 (43.3) | 401 (47.6) | 377 (47.2) | 34 (47.2) | 99 (45.2) | 32 (39.5) | 340 (39.3) |
| > 10 | 351 (35.2) | 736 (39.1) | 287 (34.1) | 309 (38.7) | 26 (36.1) | 53 (24.2) | 38 (46.9) | 374 (43.2) |
| missing | 99 (9.0) | 471 (20.0) | 7 (0.8) | 24 (2.9) | 41 (36.3) | 89 (28.9) | 51 (38.6) | 358 (29.3) |
| **Pre-referral RAS use** | | | | | | | | |
| yes | 936 (85.5) | 161 (6.8) | 781 (91.9) | 33 (4.0) | 46 (40.7) | 2 (0.7) | 109 (82.6) | 126 (10.3) |
| **Malaria test**** | | | | | | | | |
| positive (mRDT or blood slide) | 1002 (91.5) | 2245 (95.4) | 761 (89.5) | 750 (91.1) | 111 (98.2) | 281 (91.2) | 130 (98.5) | 1214 (99.3) |
| negative / not done | 93 (8.5) | 109 (4.6) | 89 (10.5) | 73 (8.9) | 2 (1.8) | 27 (8.8) | 2 (1.5) | 9 (0.7) |
| **Rainy season°°** | 707 (64.6) | 1475 (62.7) | 546 (64.2) | 518 (62.9) | 70 (62.0) | 187 (60.7) | 91 (68.9) | 770 (63.0) |
| **Drugs payable** | 529 (48.3) | 863 (36.7) | 438 (51.5) | 478 (58.1) | 77 (68.1) | 246 (79.9) | 14 (10.6) | 139 (11.4) |
| **Hospitalisation payable** | 415 (37.9) | 638 (27.1) | 458 (46.1) | 439 (53.3) | 17 (15.0) | 51 (16.6) | 6 (4.6) | 148 (12.1) |
| **Health Zone / District / LGA** | | | | | | | | |
| Kenge / Fufore / Kole | 261 (23.8) | 308 (13.1) | 261 (30.7) | 308 (37.4) | 28 (24.8) | 109 (35.4) | 43 (32.6) | 351 (28.7) |
| Kingandu / Mayo Belwa / Oyam | 189 (17.3) | 87 (3.7) | 189 (22.2) | 87 (10.6) | 69 (61.1) | 91 (29.6) | 69 (52.3) | 662 (54.1) |
| Ipamu / Song / Apac | 400 (36.5) | 428 (18.2) | 400 (47.1) | 428 (52.0) | 16 (14.2) | 108 (35.1) | 20 (15.2) | 210 (17.2) |

Number and column % of those with non-missing data, pooled, by country and by enrolment location; missing data rows are number and column %
** Severe malaria diagnosis was based on clinical assessment, diagnostic test result may or may not have been considered for the diagnosis
° WHO general danger signs
°° At time of admission; DRC: October - April; Nigeria: May - October; Uganda: April - October

**Table S3:** Provision of in-hospital vs. post-discharge ACT medication.

| | Pooled (3 countries) | | | DRC | | | Nigeria | | | Uganda | | |
|---|---|---|---|---|---|---|---|---|---|---|---|---|
| | **Community enrolments** **N = 1,091** n (%) | **RHF enrolments** **N = 2,348** n (%) | ***p* value (Chi2)** | **Community enrolments** **N = 846** n (%) | **RHF enrolments** **N = 823** n (%) | ***p* value (Chi2)** | **Community enrolments** **N = 113** n (%) | **RHF enrolments** **N = 308** n (%) | ***p* value (Chi2)** | **Community enrolments** **N = 132** n (%) | **RHF enrolments** **N = 1,217** n (%) | ***p* value (Chi2)** |
| **Provision of ACT treatment** | | | <0.001 | | | 0.021 | | | 0.200 | | | 0.002 |
| No ACT | 200 (18.3) | 316 (13.5) | | 145 (17.1) | 170 (20.7) | | 54 (47.8) | 131 (42.5) | | 1 (0.8) | 15 (1.2) | |
| ACT treatment completed at facility * | 432 (39.6) | 562 (23.9) | | 425 (50.2) | 397 (48.2) | | 0 (0.0) | 0 (0.0) | | 7 (5.3) | 165 (13.6) | |
| Started at facility, to be completed at home ◊ | 284 (26.0) | 660 (28.1) | | 251 (29.7) | 241 (29.3) | | 0 (0.0) | 7 (2.3) | | 33 (25.0) | 412 (33.9) | |
| Received prescription to buy from pharmacy ° | 155 (14.2) | 803 (34.2) | | 5 (0.6) | 9 (1.1) | | 59 (52.2) | 170 (55.2) | | 91 (68.9) | 624 (51.3) | |
| Other | 20 (1.8) | 7 (0.3) | | 20 (2.4) | 6 (0.7) | | 0 (0.0) | 0 (0.0) | | 0 (0.0) | 1 (0.1) | |
| missing | 4 (0.4) | 6 (0.3) | | 4 (0.5) | 0 (0.0) | | 0 (0.0) | 0 (0.0) | | 0 (0.0) | 6 (0.5) | |

Number and % of distribution of modalities of receiving ACT treatment, pooled, by country and enrolment location; missing data rows are number and column %
* Includes 3 children who received artesunate + mefloquine
◊ Includes 7 children who received artesunate + mefloquine and 70 observations with missing specification of type of ACT given
° Includes 1 child who received a prescription for dihydroartemisinin + piperaquine and 37 observations with missing specification of type of ACT prescribed

**Table S4:** Antimalarial treatment compliance: number of doses and total doses of injectable antimalarials administered and ACTs prescribed (subsample, post-RAS only)

| | Pooled (3 countries) | | | DRC | | | Nigeria | | | Uganda | | |
|---|---|---|---|---|---|---|---|---|---|---|---|---|
| | **Community enrolments** | **RHF enrolments** | ***p* value (Chi2)** | **Community enrolments** | **RHF enrolments** | ***p* value (Chi2)** | **Community enrolments** | **RHF enrolments** | ***p* value (Chi2)** | **Community enrolments** | **RHF enrolments** | ***p* value (Chi2)** |
| | n (%) | n (%) | | n (%) | n (%) | | n (%) | n (%) | | n (%) | n (%) | |
| **Administration of at least one dose of an inj. antimalarial[1]** | **N = 1,095** | **N = 2,354** | 0.006 | **N = 850** | **N = 823** | 0.790 | **N = 113** | **N = 308** | 0.906 | **N = 132** | **N = 1,223** | 0.942 |
| Yes | 1046 (95.5) | 2291 (97.3) | | 804 (94.6) | 776 (94.3) | | 111 (98.2) | 302 (98.1) | | 131 (99.2) | 1213 (99.2) | |
| Artesunate | 1015 (92.7) | 2234 (94.9) | 0.010 | 773 (90.9) | 736 (89.4) | 0.298 | 111 (98.2) | 298 (96.8) | 0.420 | 131 (99.2) | 1200 (98.1) | 0.353 |
| Artemether | 0 (0.0) | 8 (0.3) | 0.053 | 0 (0.0) | 3 (0.4) | 0.078 | 0 (0.0) | 4 (1.3) | 0.224 | 0 (0.0) | 1 (0.1) | 0.742 |
| Quinine | 35 (3.2) | 58 (2.5) | 0.216 | 34 (4.0) | 39 (4.7) | 0.460 | 1 (0.9) | 2 (0.7) | 0.799 | 0 (0.0) | 17 (1.4) | 0.173 |
| **Number of doses of inj. antimalarial** | **N = 1.046** | **N = 2,291** | 0.022 | **N = 804** | **N = 776** | 0.649 | **N = 111** | **N = 302** | 0.643 | **N = 131** | **N = 1,213** | 0.234 |
| < 3 | 48 (4.6) | 69 (3.0) | | 38 (4.7) | 33 (4.3) | | 10 (9.0) | 23 (7.6) | | 0 (0.0) | 13 (1.1) | |
| ≥ 3 | 998 (95.4) | 2222 (97.0) | | 766 (95.3) | 743 (95.8) | | 101 (91.0) | 279 (92.4) | | 131 (100.0) | 1200 (98.9) | |
| **In-hospital administration of at least one dose of an ACT after ≥ 3 doses of inj. treatment[2]** | **N = 1.046** | **N = 2,291** | <0.001 | **N = 804** | **N = 776** | 0.216 | **N = 111** | **N = 302** | NA | **N = 131** | **N = 1,213** | <0.001 |
| Yes | 697 (66.6) | 1188 (51.9) | | 658 (81.8) | 616 (79.4) | | 0 (0.0) | 5 (1.7) | | 39 (29.8) | 567 (46.7) | |
| **Administration / dispensing / prescription of at least one dose of an ACT after ≥ 3 doses of inj. treatment[3]** | **N = 1,046** | **N = 2,291** | <0.001 | **N = 804** | **N = 776** | 0.295 | **N = 111** | **N = 302** | 0.212 | **N = 131** | **N = 1,213** | 0.379 |
| Yes | 839 (80.2) | 1961 (85.6) | | 663 (82.5) | 624 (80.4) | | 46 (41.4) | 146 (48.3) | | 130 (99.2) | 1191 (98.2) | |
| **Number of days of ACT prescription / dispensing[4]** | **N = 373** | **N = 1,357** | 0.092 | **N = 205** | **N = 193** | 0.598 | **N = 44** | **N = 129** | 0.169 | **N = 124** | **N = 1,035** | 0.356 |
| < 3 | 7 (1.9) | 54 (4.0) | | 2 (1.0) | 1 (0.5) | | 1 (2.3) | 0 (0.0) | | 4 (3.2) | 53 (5.1) | |
| 3 | 364 (97.6) | 1300 (95.8) | | 203 (99.0) | 192 (99.5) | | 41 (93.2) | 126 (97.7) | | 120 (96.8) | 982 (94.9) | |
| >3 | 2 (0.5) | 3 (0.2) | | 0 (0.0) | 0 (0.0) | | 2 (4.6) | 3 (2.3) | | 0 (0.0) | 0 (0.0) | |
| missing | 21 (54.8) | 36 (30.6) | | 19 (8.5) | 12 (5.9) | | 2 (4.4) | 24 (15.7) | | 0 (0.0) | 0 (0.0) | |

Number and % of children receiving appropriate antimalarial treatment compliant to WHO guidelines (type of drug and number of doses), pooled, by country and enrolment location; missing data rows are number and column %

[1] More than one type of antimalarial may have been administered

[2] Compliant treatment administration

[3] Compliant treatment prescription

[4] Only includes children with a prescription / dispensing of an ACT (ALU or ASAQ) to complete treatment at home, does not include ACT treatment during hospitalisation

**Table S5:** Antimalarial dosing compliance: total doses of injectable antimalarials administered and ACTs prescribed (subsample, post-RAS only).

| | Pooled (3 countries) | | | DRC | | | Nigeria | | | Uganda | | |
|---|---|---|---|---|---|---|---|---|---|---|---|---|
| | **Community enrolments** | **RHF enrolments** | ***p* value (Chi2)** | **Community enrolments** | **RHF enrolments** | ***p* value (Chi2)** | **Community enrolments** | **RHF enrolments** | ***p* value (Chi2)** | **Community enrolments** | **RHF enrolments** | ***p* value (Chi2)** |
| | n (%) | n (%) | | n (%) | n (%) | | n (%) | n (%) | | n (%) | n (%) | |
| **Inj. antimalarial correctly dosed *** | **N = 949** | **N = 1,829** | 0.093 | **N = 798** | **N = 752** | 0.001 | **N = 71** | **N = 217** | 0.115 | **N = 80** | **N = 860** | 0.283 |
| correct | 793 (83.6) | 1481 (81.0) | | 675 (84.6) | 585 (77.8) | | 56 (78.9) | 188 (86.6) | | 62 (77.5) | 708 (82.3) | |
| under-dosed | 156 (16.4) | 348 (19.0) | | 123 (15.4) | 167 (22.2) | | 15 (21.1) | 29 (13.4) | | 18 (22.5) | 152 (17.7) | |
| *≥ 3 doses* | *112 (71.8)* | *293 (84.2)* | | *85 (69.1)* | *137 (82.0)* | | *9 (60.0)* | *12 (41.4)* | | *18 (100.0)* | *144 (94.7)* | |
| *< 3 doses* | *44 (28.2)* | *55 (15.8)* | | *38 (30.9)* | *30 (18.0)* | | *6 (40.0)* | *17 (58.6)* | | *0 (0.0)* | *8 (5.3)* | |
| missing | 101 (9.6) | 471 (20.5) | | 9 (1.1) | 26 (3.4) | | 41 (36.6) | 87 (29.0) | | 51 (38.9) | 358 (29.4) | |
| **Inj. artesunate correctly dosed** | **N = 918** | **N = 1,778** | 0.055 | **N = 767** | **N = 713** | 0.003 | **N = 71** | **N = 213** | 0.104 | **N = 80** | **N = 852** | 0.252 |
| correct | 785 (85.5) | 1469 (82.6) | | 667 (87.0) | 580 (81.4) | | 56 (78.9) | 185 (86.9) | | 62 (77.5) | 704 (82.6) | |
| under-dosed | 133 (14.5) | 309 (17.4) | | 100 (13.0) | 133 (18.7) | | 15 (21.1) | 28 (13.2) | | 18 (22.5) | 148 (17.4) | |
| missing | 97 (9.6) | 456 (20.4) | | 6 (0.8) | 23 (3.1) | | 40 (36.0) | 85 (28.5) | | 51 (38.9) | 348 (29.0) | |
| **Inj. quinine correctly dosed** | **N = 34** | **N = 49** | 0.177 | **N = 34** | **N = 38** | 0.066 | **N = 0** | **N = 2** | NA | **N = 0** | **N = 9** | NA |
| correct | 8 (23.5) | 6 (12.2) | | 8 (23.5) | 3 (7.9) | | NA (NA) | 0 (0.0) | | NA (NA) | 3 (33.3) | |
| under-dosed | 26 (76.5) | 43 (87.8) | | 26 (76.5) | 35 (92.1) | | NA (NA) | 2 (100.0) | | NA (NA) | 6 (66.7) | |
| missing | 1 (2.9) | 9 (15.5) | | 0 (0.0) | 1 (2.6) | | 1 (100.0) | 0 (0.0) | | 0 (NA) | 8 (47.1) | |
| **ACT prescription / dispensing correctly dosed °** | **N = 367** | **N = 1,339** | 0.444 | **N = 208** | **N = 193** | 0.521 | **N = 35** | **N = 113** | 0.035 | **N = 124** | **N = 1,033** | 0.854 |
| correct | 293 (79.8) | 1056 (78.9) | | 180 (86.5) | 160 (82.9) | | 9 (25.7) | 49 (43.4) | | 104 (83.9) | 847 (82.0) | |
| overdosed | 32 (8.7) | 145 (10.8) | | 6 (2.9) | 9 (4.7) | | 13 (37.1) | 20 (17.7) | | 13 (10.5) | 116 (11.2) | |
| under-dosed | 42 (11.4) | 138 (10.3) | | 22 (10.6) | 24 (12.4) | | 13 (37.1) | 44 (38.9) | | 7 (5.7) | 70 (6.8) | |
| *= 3 days* | *35 (83.3)* | *86 (62.3)* | | *20 (90.9)* | *23 (95.8)* | | *11 (84.6)* | *44 (100.0)* | | *4 (57.1)* | *19 (27.1)* | |
| *< 3 days* | *6 (14.3)* | *52 (37.7)* | | *2 (9.1)* | *1 (4.2)* | | *1 (7.7)* | *0 (0.0)* | | *3 (42.9)* | *51 (72.9)* | |
| missing | 458 (55.5) | 615 (31.5) | | 440 (67.9) | 407 (67.8) | | 11 (23.9) | 40 (26.1) | | 7 (5.3) | 168 (14.0) | |
| **ALU prescription / dispensing correctly dosed °** | **N = 190** | **N = 1,161** | 0.003 | **N = 31** | **N = 15** | 0.913 | **N = 35** | **N = 113** | 0.035 | **N = 124** | **N = 1,033** | 0.854 |
| correct | 129 (67.9) | 904 (77.9) | | 16 (51.6) | 8 (53.3) | | 9 (25.7) | 49 (43.4) | | 104 (83.9) | 847 (82.0) | |
| overdosed | 26 (13.7) | 136 (11.7) | | 0 (0.0) | 0 (0.0) | | 13 (37.1) | 20 (17.7) | | 13 (10.5) | 116 (11.2) | |
| under-dosed | 35 (18.4) | 121 (10.4) | | 15 (48.4) | 7 (46.7) | | 13 (37.1) | 44 (38.9) | | 7 (5.7) | 70 (6.8) | |
| missing | 18 (8.7) | 208 (15.2) | | 8 (20.5) | 15 (50.0) | | 3 (7.9) | 25 (18.1) | | 7 (5.3) | 168 (14.0) | |
| **ASAQ prescription / dispensing correctly dosed °** | **N = 177** | **N = 178** | 0.074 | **N = 177** | **N = 178** | 0.074 | **N = 0** | **N = 0** | NA | **N = 0** | **N = 0** | NA |
| correct | 164 (92.7) | 152 (85.4) | | 164 (92.7) | 152 (85.4) | | NA (NA) | NA (NA) | | NA (NA) | NA (NA) | |
| overdosed | 6 (3.4) | 9 (5.1) | | 6 (3.4) | 9 (5.1) | | NA (NA) | NA (NA) | | NA (NA) | NA (NA) | |
| under-dosed | 7 (4.0) | 17 (9.6) | | 7 (4.0) | 17 (9.6) | | NA (NA) | NA (NA) | | NA (NA) | NA (NA) | |

Number and % of children receiving antimalarial treatment compliant to WHO guidelines (total dosis), pooled, by country and enrolment location; missing data rows are number and column %
* defined as min. 3 doses of either artesunate (total 9 mg / kg bw), artemether (total 6.4 mg / kg bw) or quinine (total 40 mg / kg bw), per WHO 2015 treatment guideline
° defined as: artemether-lumefantrine: 4 months - 3 years: 3x2x20/120 mg; > 3 years: 3x2x40/240 mg; artesunate-amodiaquine: < 1 year: 3x25/67.5 mg; ≥ 1 year: 3x 75/135 mg (WHO 2015 treatment guideline)

**Supplementary materials S6:** Assessment of correct dosing of injectable antimalarials and ACTs.

Correct dosing of injectable antimalarials (intravenous (IV) or intramuscular (IM)) meant the administration of at least three doses at the WHO recommended dose, i.e. a minimal total dose of artesunate of at least 9 mg/kg body weight, 6.4 mg/kg body weight for injectable artemether and 40 mg/kg body weight for quinine.

Correct dosing was of ACTs was defined as a prescription for 3 days of ALU at the recommended dose (total dose of 120 mg artemether + 720 mg lumefantrine for age up to three years, 240 mg artemether + 1,440 mg lumefantrine for older than three years) or ASAQ (total dose of 75 mg artesunate + 202.5 mg amodiaquine for children below 1 year of age, 150 mg artesunate + 405 mg amodiaquine for children older than one year) constituted correct dosing. Since weight was missing for a considerable fraction of patients in the subsample (16.5%), the correctness of ACT dosing was alternatively estimated based on patient age, as described previously (5, 6). The dosing assessment was limited to ACT prescriptions issued at discharge and did not include in-hospital ACT therapy due to incomplete data collection.

### References


1. World Health Organization. Guidelines for the treatment of malaria. 3rd ed: World Health Organization; 2015.
2. République Démocratique du Congo - Ministère de la Santé Publique SG. Directives Nationales de Prise en Charge du Paludisme. 2016 [Available from: https://www.severemalaria.org/sites/mmv-smo/files/content/attachments/2017-11-20/DRC%20DIRECTIVES%20NATIONALE%20DE%20PEC%20DU%20PALUDISME%202017.pdf.
3. Federal Ministry of Health Nigeria. National Guidelines for Diagnosis and Treatment of Malaria. 3rd ed. Abuja Nigeria.2015.
4. Republic of Uganda - Ministry of Health. Uganda Clinical Guidelines. 2016 [Available from: https://health.go.ug/sites/default/files/Uganda%20Clinical%20Guidelines%202016_FINAL.pdf.
5. Taylor WR, Terlouw DJ, Olliaro PL, White NJ, Brasseur P. Use of weight-for-age-data to optimize tablet strength and dosing regimens for a new fixed-dose artesunate- amodiaquine combination for treating falciparum malaria. Bulletin of the World Health Organization. 2006:10.
6. World Health Organization. Guidelines for the treatment of malaria. 1st ed: World Health Organization; 2006.
